# The effects of COVID-19 on child mental and social health: biannual assessments up to April 2022 in a clinical and two general population samples

**DOI:** 10.1101/2022.09.08.22279670

**Authors:** Josjan Zijlmans, Jacintha M. Tieskens, Hedy A. van Oers, Hekmat Alrouh, Michiel A.J. Luijten, Rowdy de Groot, Daniël van der Doelen, Helen Klip, Rikkert M. van der Lans, Ronald de Meyer, Malindi van der Mheen, I. Hyun Ruisch, Germie van den Berg, Hilgo Bruining, Jan Buitelaar, Rachel van der Rijken, Pieter J. Hoekstra, Marloes Kleinjan, Ramón J.L. Lindauer, Kim J. Oostrom, Wouter Staal, Robert Vermeiren, Ronald Cornet, Lotte Haverman, Arne Popma, Meike Bartels, Tinca J. C. Polderman

## Abstract

**Background:** The COVID-19 pandemic has had an acute impact on child mental and social health, but long-term effects are still unclear. We examined how child mental health has developed since the start of the COVID-19 pandemic up to two years into the pandemic (April 2022).

**Methods:** We included children (age 8-18) from two general population samples (N = 222-1,333 per measurement and N = 2,401-13,362 for pre-covid data) and one clinical sample receiving psychiatric care (N = 334-748). Behavioral questionnaire data were assessed five times from April 2020 till April 2022 and pre-pandemic data were available for both general population samples. We collected parent-reported data on internalizing and externalizing problems with the Brief Problem Monitor and self-reported data on Anxiety, Depressive symptoms, Sleep-related impairments, Anger, Global health, and Peer relations with the Patient-Reported Outcomes Measurement Information System (PROMIS®).

**Results:** In all samples, parents reported overall increased internalizing problems, but no increases in externalizing problems, in their children. Children from the general population self-reported increased mental health problems from before to during the pandemic on all six PROMIS domains, with generally worst scores in April 2021, and scores improving towards April 2022 but not to pre-pandemic norms. Children from the clinical sample reported increased mental health problems throughout the pandemic, with generally worst scores in April 2021 or April 2022 and no improvement. We found evidence of minor age effects and no sex effects.

**Conclusions:** Child mental health in the general population has deteriorated during the first phase of the COVID-19 pandemic, has improved since April 2021, but has not yet returned to pre-pandemic levels. Children in psychiatric care show worsening of mental health problems during the pandemic, which has not improved since. Changes in child mental health should be monitored comprehensively to inform health care and policy.

## Introduction

The COVID-19 pandemic and subsequent restrictions such as social distancing, the closing of schools, and even total lockdowns severely disrupted people’s lives. Children and adolescents (hereafter referred to as children) are less susceptible to physical symptoms of COVID-19 infections (Viner et al., 2021). However, they may be more prone to negative indirect effects of the pandemic such as lockdowns and other restrictions that could disrupt their social networks and may be at risk of developing mental health problems (Danese et al., 2020).

Research into the effects of disasters and emergencies, such as pandemics, wars, and natural disasters, has shown that children’s resilience to traumatic events varies greatly. For instance, increases in mental health problems in response to traumatic experiences rapidly decrease to normal levels in some children, while for other children the consequences can be long-lasting (Masten & Motti-Stefanidi, 2020; Sonuga-Barke & Fearon, 2021). During the pandemic, children with pre-existing mental health problems may be particularly at risk of sustained negative mental health effects, as their pre-existing mental health problems might exacerbate and as their mental healthcare was often interrupted or altered (Hoffmann & Duffy, 2021; Witt et al., 2020). In addition, some studies have reported that girls and older children (13-15 compared to 6-12 years old) may be more strongly impacted by the pandemic (Panchal et al., 2021), but as pre-pandemic data are often missing, it is hard to conclude whether these are COVID-specific or general effects.

Indeed, systematic reviews and meta-analyses conclude that the mental health of children worldwide was negatively affected during the first year of the COVID-19 pandemic (Ma et al., 2021; Panchal et al., 2021; Samji et al., 2022; Zolopa et al., 2022), but these interpretations were mostly based upon cross-sectional studies performed during the pandemic. Prospective studies on child mental health during the pandemic that include pre-pandemic measurements are scarce but do seem to confirm that predominantly affective problems such as depressive, anxious, and stress symptoms worsened in the first months after the onset of the COVID-19 pandemic. This seems to be the case for children from the general population (Barendse et al., 2022; Bignardi et al., 2021; de France et al., 2022; Fischer et al., 2022; Luijten, van Muilekom, et al., 2021; van der Velden et al., 2022) as well as for children with pre-existing mental health problems (Breaux et al., 2021; Fischer et al., 2022). On the contrary, some other studies challenge the mental burden of the COVID-19 pandemic in children and report small or no significant differences in mental health problems (Bouter et al., 2022; Burdzovic Andreas & Brunborg, 2021).

To date, no studies have reported on the effects of the pandemic on child mental health past early 2021 (the first year of the pandemic). However, to be able to provide adequate measures and counseling, it is important for policy makers and health care professionals to know whether problems have exacerbated, stabilized, or normalized since then.

In this study, we aimed to quantify how child mental and social health (hereafter referred to as mental health) has developed since the start of the COVID-19 pandemic up to two years into the pandemic (April 2022). We investigated whether changes are different for children with pre-existing mental health problems compared to children from the general population, and whether sex and age impact COVID-related mental health changes. We systematically examined the mental health of children from the general population and that of those in psychiatric care using data from before the COVID-19 pandemic and as assessed over the course of the pandemic biannually from April 2020 till April 2022. We collected parent-reported and self-reported outcome measures on multiple domains of mental health, including internalizing and externalizing problems.

## Methods

### Participants

The Dutch consortium Child and Adolescent Mental Health and WellBeing in times of the COVID-19 pandemic (CAMHWB-19) was initiated to assess the impact of the COVID-19 pandemic on the mental health and wellbeing of children and adolescent in the Netherlands. It comprises (parents of) children aged 8-18 from two general population samples and one clinical sample. We previously reported on the first two pandemic measurements of this study (Fischer et al., 2022). The two population samples are 1) The Netherlands Twin Register (NTR; Ligthart et al., 2019), a twin cohort that has collected data over the past 35 years in the general Dutch population; 2) The KLIK group, which aimed to collect samples representative of the Dutch population using an online panel agency. The clinical sample is 3) DREAMS (Dutch Research in child and Adolescent Mental health), a collaboration between four academic child and adolescent psychiatry centers (Amsterdam, Groningen, Leiden, Nijmegen) in the Netherlands that has obtained information from children and their parents receiving psychiatric care particularly for this study. Sample sizes varied between 222 and 1,333 for each measurement during the COVID pandemic and are up to 13,362 before the COVID pandemic.

Collaborating parties received approval for data collection by the appropriate ethics committees and all children and parents provided informed consent. The studies were conducted in line with the ethical standards stated in the 1964 Declaration of Helsinki and its later amendments.

### Procedure

Data were collected at five moments in time after the start of the pandemic, approximately once every six months. At each moment, both new and recurrent participants were invited to participate. To prevent within-subject effects biasing the results, in all samples we randomly selected one measurement occasion for each individual participant. Pre-pandemic data were available for the two general population samples, but not for the clinical sample. Table 1 presents an overview of the samples and data that were used for the analyses.

**Table 1.** Sociodemographic Characteristics of the Samples

| Cohort |  | 0<br>pre-pandemic | 1<br>Apr/May 2020 | 2<br>Nov/Dec 2020 | 3<br>Mar/Apr 2021 | 4<br>Nov/Dec 2021 | 5<br>Mar/Apr 2022 |
| --- | --- | --- | --- | --- | --- | --- | --- |
| NTR | N total | 13362 | 1791 | 536 | 714 | 855 | 883 |
|  | N after random selection* | 13362 | 1333 | 222 | 347 | 426 | 458 |
|  | Males | 49.5% | 49.3% | 49.6% | 55.0% | 47.4% | 49.6% |
|  | M age (SD) | 11.6 (1.3) | 10.7 (1.8) | 10.3 (2.0) | 10.1 (2.1) | 10.9 (2.2) | 11.1 (1.9) |
|  | Country of birth parents (both Dutch) | 88.8% | 83.4% | 83.3% | 73.8% | 83.8% | 81.2% |
|  | Educational level parents low | 20.5% | 3.9% | 1.4% | 5.2% | 0.0% | 4.8% |
|  | Educational level parents intermediate | 38.2% | 29.9% | 38.6% | 31.0% | 30.0% | 14.3% |
|  | Educational level parents high | 41.3% | 66.2% | 60.0% | 63.8% | 70.0% | 81.0% |
| KLIK | N total | 2401 | 856 | 939 | 909 | 828 | 893 |
|  | N after random selection* | 2401 | 486 | 440 | 413 | 414 | 529 |
|  | Males | 50.3% | 47.9% | 50.2% | 52.1% | 50.5% | 54.1% |
|  | M age (SD) | 13.1 (3.1) | 13.5 (2.9) | 13.8 (3.2) | 13.6 (3.3) | 13.6 (3.1) | 13.4 (3.1) |
|  | Country of birth parents (both Dutch) † | 79.8% | 86.6% | 88.9% | 88.5% | 88.5% | 89.7% |
|  | Educational level parents low † | 12.8% | 9.1% | 9.4% | 9.8% | 7.5% | 7.8% |
|  | Educational level parents intermediate | 48.2% | 52.0% | 45.3% | 48.4% | 45.6% | 48.7% |
|  | Educational level parents high | 38.9% | 38.9% | 45.3% | 41.8% | 46.9% | 43.5% |
| DREAMS | N total | - | 500 | 892 | 661 | 632 | 450 |
|  | N after random selection* | - | 453 | 748 | 505 | 482 | 334 |
|  | Males | - | 54.3% | 53.5% | 59.6% | 53.7% | 47.0% |
|  | M age (SD) | - | 13.3 (3.1) | 13.6 (2.9) | 13.0 (2.9) | 13.2 (3.0) | 13.3 (2.9) |
|  | Country of birth parents (both Dutch) | - | 84.3% | 88.4% | 86.2% | 85.4% | 84.5% |
|  | Educational level parents low | - | 4.8% | 5.6% | 6.3% | 6.9% | 6.5% |
|  | Educational level parents intermediate | - | 40.3% | 45.6% | 42.4% | 44.4% | 38.1% |
|  | Educational level parents high | - | 54.9% | 48.8% | 51.3% | 48.7% | 55.4% |

Note. Statistics represent the samples *after* random selection of a single measurement moment for each participant. N = number of participants, M = mean, SD = standard deviation.
\* = Number of participants after random selection of one measurement for each individual participant.
† = Sample size is smaller as data were not collected in norm studies for PROMIS anxiety and depressive symptoms.

Data were collected in April/May 2020 (measurement 1), November/December 2020 (measurement 2), March/April 2021 (measurement 3), November/December 2021 (measurement 4), and March/April 2022 (measurement 5). Measurement 1 was during the first peak of the pandemic when the first lockdown was set in The Netherlands and all schools were closed. Measurement 2 was during a partial lockdown with schools partially reopened. Measurement 3 was also during partial lockdown with the addition of a nighttime curfew. Measurement 4 was again during (partial) lockdown, schools were still open but right after our data collection schools closed again (on December 14^th^). Measurement 5 was during a relaxation of most of the restrictions. Schools were open, people were allowed to work from the office, and facemasks were no longer mandatory in most public spaces. See Figure 1 for an overview of the data collection and Dutch COVID restrictions at the time.

**Figure 1.**
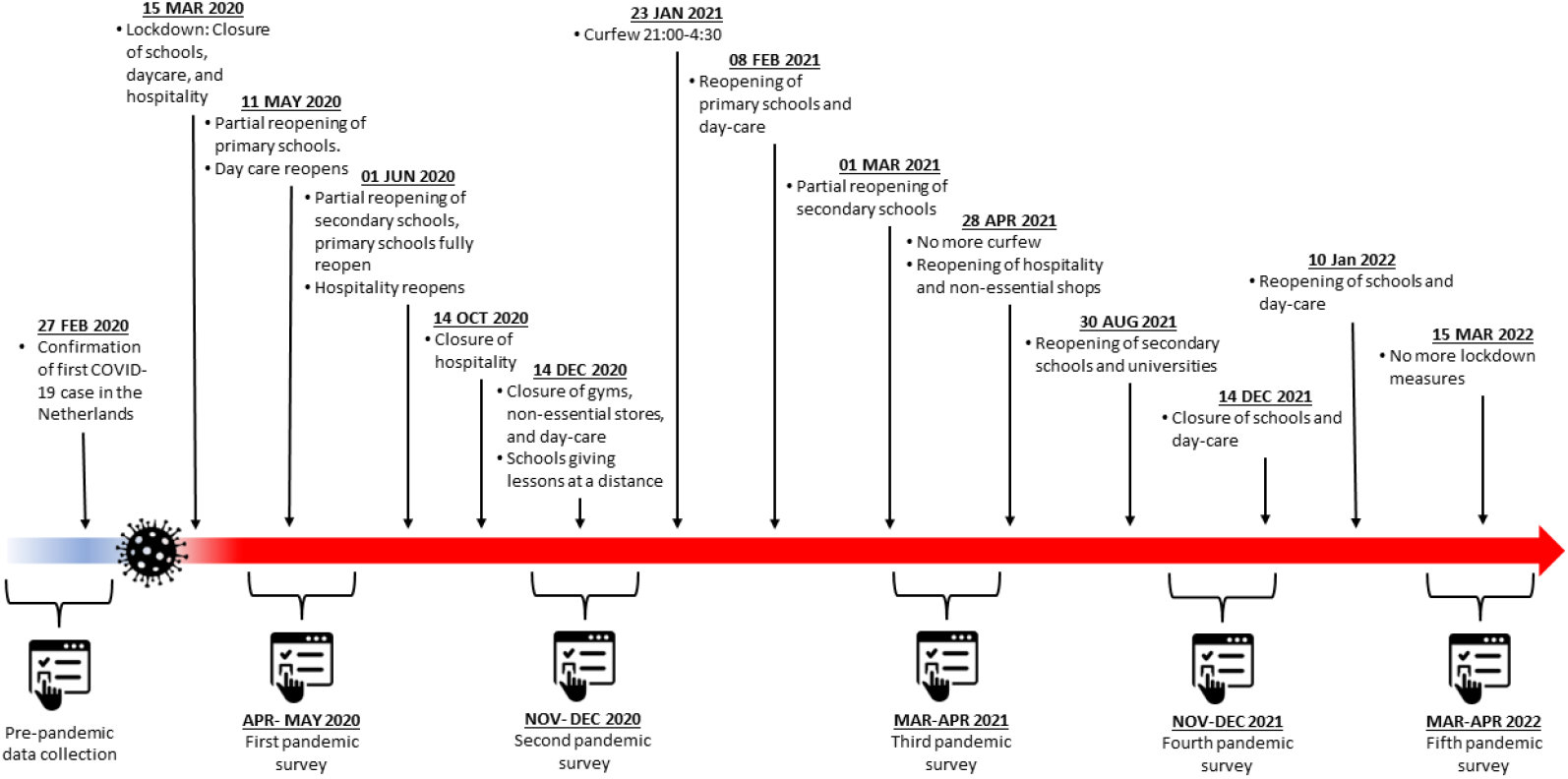
Timeline of COVID-19 restrictions in the Netherlands from April 2020 to April 2022

NTR invited parents of twins via email for a regular survey (measurement 2) and for four COVID specific additional surveys. Pre-pandemic data were available from previous standard cohort data collections. Response rates were chronologically 45%, 38%, 31%, 31%, 19%, 14%.

KLIK invited children and their parents via an independent, online survey panel (PanelInzicht). (Luijten, van Litsenburg, et al., 2021). Data were collected through a research website of the KLIK portal (www.corona.hetklikt.nu). Pre-pandemic data were available from previous validation studies of the PROMIS measures. The pre-pandemic group was representative of the Dutch general population within 2.5% on most key demographics (age, gender, ethnicity, region, and educational level) compared to population numbers in 2017. Samples drawn during the pandemic are similar but include slightly more children of whom both parents were born in the Netherlands and slightly fewer children from low-income families (see table 1).

DREAMS invited children receiving psychiatric care and their parents via e-mail through their respective psychiatric centers. Data were collected via the same research portal as the KLIK group employed. Response rates were respectively 9%, 10%, 11%, 10%, and 9%.

### Measures

#### Socio-demographic information

To describe the samples, we gathered data on the country of birth and educational level of the parents. Country of birth was operationalized as both parents being born in the Netherlands (yes/no). Educational level was operationalized as the highest education among both parents (Low = primary, lower vocational education, lower and middle general secondary education; Intermediate = middle vocational education, higher secondary education, pre-university education; High = higher vocational education, university).

#### Parent-Reported Outcomes

For parental reports in NTR and DREAMS, we employed the Brief Problem Monitor of the Achenbach System of Empirical Based Assessment (ASEBA-BPM). The BPM (Achenbach et al., 2011) is a shortened version of the Child Behavior Checklist (CBCL/6-18 years; (Achenbach & Rescorla L A, 2001). It assesses behavioral and emotional problems in children as reported by their parents. Items are rated on a three-point Likert-scale, where parents rate if a statement applies to their child (0 = ‘not true’, 1 = ‘somewhat true’, 2 = ‘very true’). In line with the BPM manual, we coded missing items on the BPM as zero. If more than 20% of items were missing, we excluded the participant from the BPM analysis. The BPM yields an internalizing score calculated from six items and an externalizing score. The externalizing score usually is calculated from seven items, but because one item pertains to behavior at school and data were also collected when children did not go to school, we excluded this item. The six remaining items were weighted so that the sum score has the same range as the normal scoring to allow comparison to other studies.

#### Child-Reported Outcomes

For child self-reports in KLIK and DREAMS, we employed the *Patient-Reported Outcomes Measurement Information System (PROMIS®)*. Six measures of the Dutch-Flemish PROMIS® were used to assess self-reported Anxiety v2.0 (Irwin et al., 2010), Depressive Symptoms v2.0 (Irwin et al., 2010), Anger v2.0 (Irwin et al., 2012), Sleep-related impairment v1.0 (Forrest et al., 2018), Global health v1.0 (Forrest et al., 2014), and Peer Relationships v2.0 (DeWalt et al., 2013). All instruments except Anger and Global Health were administered as Computerized Adaptive Tests (CAT), where items are selected based on responses to previously completed items, resulting in reliable scores with fewer items (Cella et al., 2007). PROMIS measures use a 7-day recall period, and most items are scored on a five-point Likert scale ranging from ‘never’ to ‘(almost) always’. Total scores are calculated by transforming item scores into T-scores ranging from 0 to 100 with a mean of 50 and standard deviation of 10 in the original calibration sample (Irwin et al., 2010). The US item parameters were used in the CAT algorithm and T-score calculations, as by PROMIS convention. The PROMIS pediatric item banks and scales have previously been validated in the Dutch population (Klaufus et al., 2021; Luijten et al., 2022; Luijten, van Litsenburg, et al., 2021; Peersmann et al., 2022; van Muilekom et al., 2021)

#### Data analysis

Statistical analyses were performed in IBM SPSS Statistics 28. Within each sample and for each outcome variable, we performed analyses of covariance (ANCOVA) to test whether mental health problems were different over the course of the pandemic. In all analyses, age and sex were included as covariates and we tested for interaction effects between time and sex and time and age. For the latter interaction, we split age into two groups: children below the age of 12 years old and of age 12 and higher. We performed post-hoc Least Significant Differences tests to compare individual measurement moments within each sample. For the BPM measures, we report differences in scores expressed as estimated marginal means (EMM) of Z-scores standardized to pre-pandemic data of the NTR for ease of interpretation (see Table 2). To facilitate comparison to other (international) studies, sum scores are presented in Table S1. Likewise, for the PROMIS measures, we report differences in scores expressed as EMMs of Z-scores standardized to pre-pandemic norm scores for KLIK in Table 2, and for international comparison, T-scores based on the original (United States) calibration sample are reported in Table S2.

**Table 2.**
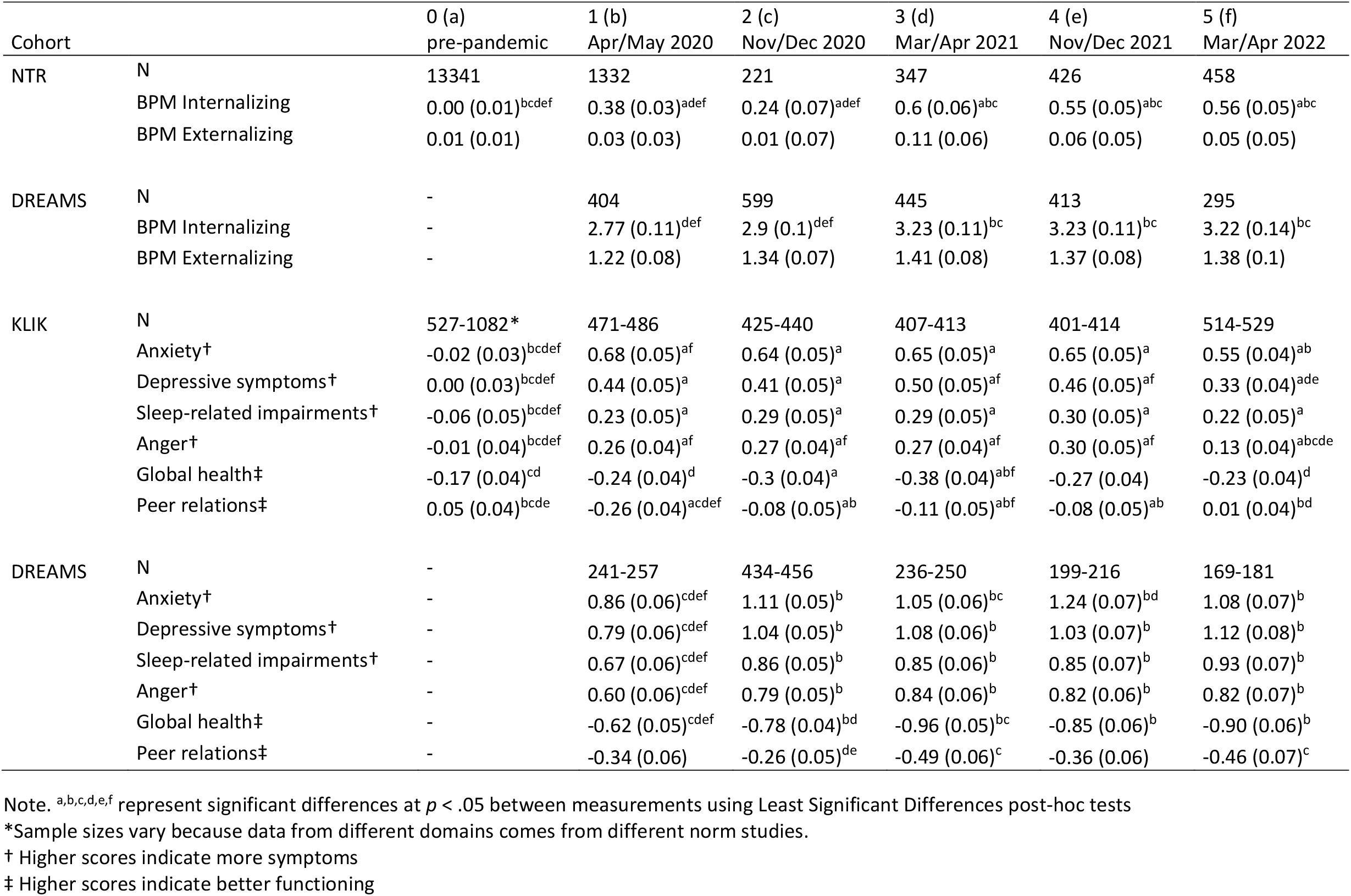
BPM and PROMIS standardized estimated marginal means (EMM), standard errors, and comparisons between measurement points

Finally, we report the proportions of children who scored outside of the normal range on the BPM internalizing and externalizing scales based on rater and sex specific T-scores (T > 65) in Table S1 and the proportion of children who scored outside of the normal range on the PROMIS scales in Table S3.

## Results

### Parent-reported outcomes (Brief Problem Monitor)

Table 2 presents results for the BPM outcome measures of NTR and DREAMS, and Figure 2 illustrates the EMMs of the general population sample and clinical sample over time, represented as standard deviations from pre-covid NTR norm scores.

**Figure 2.**
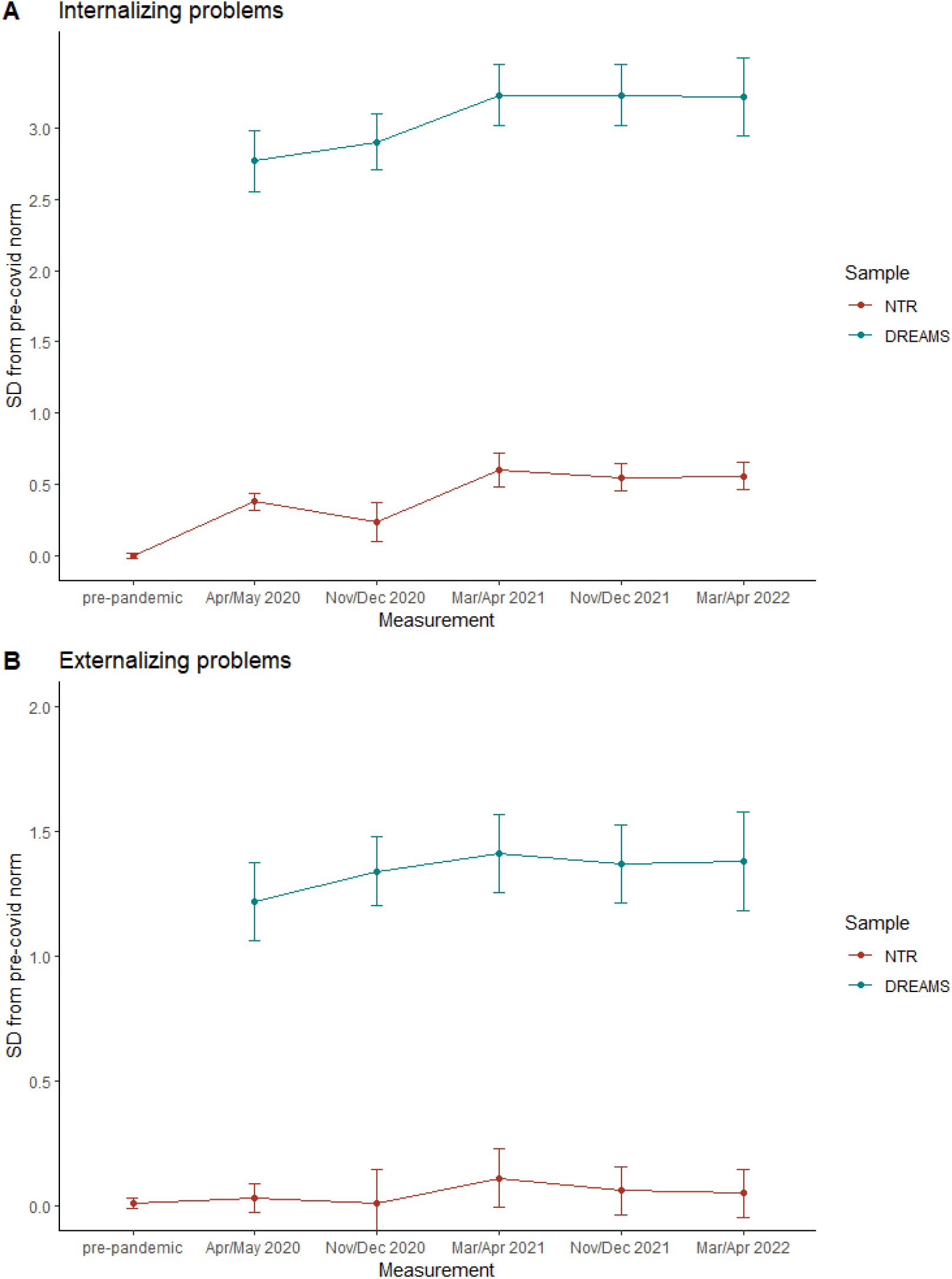
BPM standardized estimated marginal means (EMM) and confidence intervals Note. SD = standard deviation

In the general population sample of NTR, internalizing problems differed significantly between measurements (F = 80.61, p < .001), with lowest scores during the pre-COVID measurement and highest scores during the 3^rd^ COVID measurement (Mar/Apr 2021). The final COVID measurement (Mar/Apr 2022) had significantly higher scores than the pre-pandemic measurement and did not differ from the measurement with the highest scores. There was a significant interaction between time and age (F = 4.25, p < .001), where younger children had a steeper increase in problems during the 1^st^ COVID measurement (Apr/May 2020) and then returned to the same pattern as the older children. There was no interaction between time and sex.

Externalizing problems did not significantly differ between measurements. There was a significant interaction between time and age (F = 3.28, p < .01), where younger children varied little between measurements and older children more so, but both groups did not change from pre-covid to the final measurement. There were no interaction effects between time and sex.

In the clinical sample of DREAMS (note that there were no pre-pandemic data available for this sample), internalizing problems differed significantly between measurements (F = 3.86, p < .01), with lowest problem scores during the 1^st^ COVID measurement (Apr/May 2020) and highest scores during the 3^rd^ COVID measurement (Mar/Apr 2021). The final COVID measurement (Mar/Apr 2022) had significantly higher scores than the 1^st^ COVID measurement and did not differ from the measurement with the highest scores. There were no interaction effects between time and age or time and sex.

Externalizing problems did not significantly differ between measurements. There were no interaction effects between time and age or time and sex.

### Child-reported outcomes (PROMIS)

Table 2 presents results for the PROMIS outcome measures of the KLIK and DREAMS samples, and Figure 3 illustrates the EMMs of the general population sample and clinical sample over time, represented as standard deviations from pre-covid norm scores. Table S2 shows the EMMs of the U.S. calibrated T-scores for international comparison.

**Figure 3.**
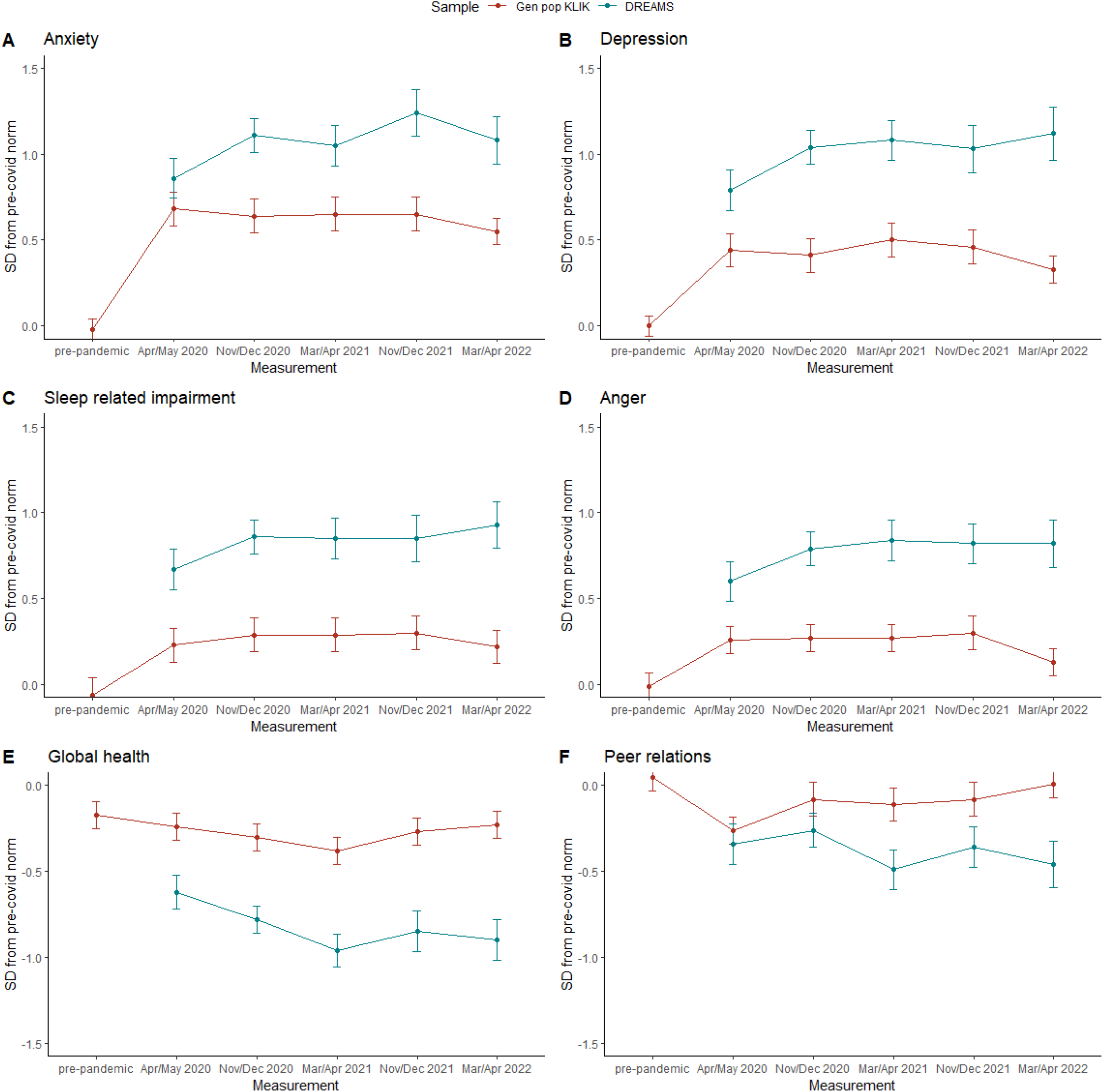
PROMIS standardized estimated marginal means (EMM) and confidence intervals Note. For figures A through D, higher scores indicate more symptoms. For figures E and F, higher scores indicate better functioning. SD = standard deviation

In the KLIK sample, levels of anxiety differed significantly between measurements (F = 74.69, p < .001), with lowest problem scores during the pre-pandemic measurement and highest scores during the 1^st^ COVID measurement (Apr/May 2020). The final COVID measurement (Mar/Apr 2022) had significantly higher scores than the pre-pandemic measurement and significantly lower scores than the measurement with the highest scores.

Levels of depression differed significantly between measurements (F = 33.13, p < .001), with lowest scores during the pre-pandemic measurement and highest scores during the 3^rd^ COVID measurement (Mar/Apr 2021). The final COVID measurement (Mar/Apr 2022) had significantly higher scores than the pre-pandemic measurement and significantly lower scores than the measurement with the highest scores.

Sleep-related impairments differed significantly between measurements (F = 9.23, p < .001), with lowest scores during the pre-pandemic measurement and highest scores during the 3^rd^ COVID measurement (Mar/Apr 2021). The final COVID measurement (Mar/Apr 2022) had significantly higher scores than the pre-pandemic measurement and did not differ from the measurement with the highest scores. There was a significant interaction between time and age (F = 2.82, p < .05), where younger children showed a larger increase in impairment from the pre-pandemic measurement to the 1^st^ COVID measurement (Apr/May 2020).

Anger differed significantly between measurements (F = 8.95, p < .001), with lowest scores during the pre-pandemic measurement and highest scores during the 3^rd^ COVID measurement (Mar/Apr 2021). The final COVID measurement (Mar/Apr 2022) had significantly higher scores than the pre-pandemic measurement and significantly lower scores than the measurement with the highest scores.

Global health differed significantly between measurements (F = 3.10, p < .01), with best scores during the pre-pandemic measurement and worst scores during the 3^rd^ COVID measurement (Mar/Apr 2021). The final COVID measurement (Mar/Apr 2022) did not differ from the pre-pandemic measurement and had significantly better scores than the measurement with the worst scores. There was a significant interaction between time and age (F = 2.34, p < .05), where older children improved after the 3^rd^ COVID measurement (Mar/Apr 2021), but younger children did not.

Peer relations differed significantly between measurements (F = 6.57, p < .001), with best scores during the pre-pandemic measurement and worst scores during the 1^st^ COVID measurement (Apr/May 2020). The final COVID measurement (Mar/Apr 2022) did not differ from the pre-pandemic measurement and had significantly better scores than the measurement with the worst scores.

Apart from the interactions for sleep-related impairments and global health, none of the interactions between time and age and time and sex on PROMIS outcomes were significant.

In the DREAMS sample (note that there were no pre-pandemic data available for this sample), levels of anxiety differed significantly between measurements (F = 5.32, p < 0.001), with lowest scores during the 1^st^ COVID measurement (Apr/May 2020) and highest scores during the 4^th^ COVID measurement (Nov/Dec 2021). The final COVID measurement (Mar/Apr 2022) had significantly higher scores than the 1^st^ COVID measurement and did not differ from the measurement with the highest scores.

Levels of depression differed significantly between measurements (F = 4.34, p < 0.01), with lowest scores during the 1^st^ COVID measurement (Apr/May 2020) and highest scores during the final COVID measurement (Mar/Apr 2022). The final COVID measurement had significantly higher scores than the 1^st^ COVID measurement.

Sleep-related impairments differed significantly between measurements (F = 2.64, p < .05), with lowest scores during the 1^st^ COVID measurement (Apr/May 2020) and highest scores during the final COVID measurement (Mar/Apr 2022). The final COVID measurement had significantly higher scores than the 1^st^ COVID measurement.

Anger differed significantly between measurements (F = 3.40, p < .01), with lowest scores during the 1^st^ COVID measurement (Apr/May 2020) and highest scores during the 3^rd^ COVID measurement (Mar/Apr 2021). The final COVID measurement (Mar/Apr 2022) had significantly higher scores than the 1^st^ COVID measurement and did not differ from the measurement with the highest scores.

Global health differed significantly between measurements (F = 6.20, p < .001), with best scores during the 1^st^ COVID measurement (Apr/May 2020) and worst scores during the 3^rd^ COVID measurement (Mar/Apr 2021). The final COVID measurement (Mar/Apr 2022) had significantly worse scores than the 1^st^ COVID measurement and did not differ from the measurement with the worst scores.

Peer relations differed significantly between measurements (F = 2.90, p < .05), with best scores during the 2^nd^ COVID measurement (Nov/Dec 2020) and worst scores during the 3^rd^ COVID measurement (Mar/Apr 2021). The final COVID measurement (Mar/Apr 2022) did not differ from the 1^st^ COVID measurement and had significantly better scores than the measurement with the worst scores.

None of the interactions between time and age and time and sex were significant.

## Discussion

In this study we assessed parent-reported and self-reported child mental and social health at five cross-sectional measurements during the pandemic in two samples of the general population and one clinical sample in the Netherlands (age 8 to 18).

### General population samples

In the NTR sample, for which we have parent-reported data, no differences were observed in externalizing problems from pre-pandemic to pandemic measurements nor over the course of the pandemic. However, parents did report higher internalizing problems of their children during the pandemic compared to before the pandemic, and in April 2022 this had not yet normalized. In the KLIK sample for which we have self-reported data, we observed significant increases in mental health problems from before the pandemic to the first measurement during the pandemic (April 2020) on all six mental health domains. In line with previous research, we found the largest increases in anxiety and depression (Barendse et al., 2022; Bignardi et al., 2021; de France et al., 2022). After this initial deterioration the problems stabilized and, in most domains, started to normalize. All measures except for sleep-related impairment were significantly better in April 2022 compared to the worst measurement during COVID (either April 2020 or April 2021). However, on all domains, except for peer relations, children still reported significantly more problems in April 2022 than before the pandemic. This is a slightly different pattern from the parent-report of the NTR, where no normalization was visible. Likely, these differences are partly due to the difference in samples, instruments, and raters. Future measurements should assess whether the normalization follows through or if additional intervention is needed.

### Clinical sample

In the clinical sample, we found self-reported mental health to worsen throughout the pandemic on all mental health domains except peer relations, which remained stable. Like the self-reports of the general population, differences were largest for anxiety and depression. Contrary to the general population, by April 2022 none of the measures had significantly improved compared to the worst measurement during COVID and on all mental health domains (except peer relations) children reported significantly more problems than during the first COVID measurement in April 2020. The parent-reported outcomes regarding internalizing problems are similar, although the effects seem delayed; parents reported increased internalizing problems from April 2021 onwards, but not before. In previous work we discussed that internalizing problems may be less readily noticed by parents or may be perceived as less problematic (Fischer et al., 2022). However, the current data suggest that parents do notice internalizing problems of their children, although in a later stage of the pandemic than children themselves. Like in the general population, parents of children from the clinical sample did not report differences in externalizing problems throughout the pandemic. Our results indicate that children in psychiatric care recuperate more slowly from the effects of the pandemic than children from the general population. Possibly, this is because although pandemic measures have been lifted, it takes time to alleviate the effects of disrupted care. Mental health care systems themselves are still recuperating and are now treating children with increased problems compared to before the pandemic. Likely, this has resulted in a higher burden for mental health care.

### Sex and age effects

Whereas other studies have suggested that girls and older children may be more vulnerable to effects of the pandemic (Panchal et al., 2021), we did not find evidence that problems increased more in girls or adolescents throughout the pandemic. As most published studies reported on single cross-sectional measurements, we suspect many reported effects of age and sex to be non-specific to the COVID-19 pandemic. Nonetheless, some longitudinal studies have observed more pronounced changes in girls. For example, Magson et al. (2021) reported greater increases in depressive and anxious symptoms in adolescent girls compared to adolescent boys (age 13 to 16). It is possible that our broader age range makes it harder to detect sex effects and that more focused studies are needed to reveal these. We did find minor differences between age groups in mental health progression throughout the pandemic, where younger children showed faster increases in sleep-related problems and slower recuperation of global health. However, the effects are small, and no consistent pattern emerges from the different measures. As there is much variability in how children have reacted to the pandemic, it would be worthwhile for future studies to investigate more specific determinants of better and worse outcomes (e.g., certain mental health problems or family circumstances). This may help policy makers adjust their approach accordingly.

### Strengths and limitations

Our study has several strengths. First, by assessing mental health in three large samples at multiple, independent, cross-sectional measurements before and since the pandemic, we avoided contamination of pandemic effects with treatment effects, developmental effects, and other time effects. For example, as children age, they are expected to report different mental health problems and as children go through treatment on average they will improve, but many longitudinal designs do not take such effects into account. Ideal designs include longitudinal measurements *within* cohorts that can be compared between *different* comparable cohorts, but such designs are often impossible because multiple pre-covid measurements as well as multiple pandemic measurements must be available in similar but different cohorts. See Burdzovic Andreas & Brunborg (2021) and Elmer et al. (2020) for good examples of such designs. Second, where most studies employ convenience samples, we included one long-term cohort sample, one representative sample of the general population, and one sample of children receiving psychiatric care, reducing selection bias, and increasing generalizability. Finally, we used well-validated measures to assess mental health problems whereas a recent review concluded that fewer than 15% of available studies used validated instruments (Samji et al., 2022).

Our study also has several limitations. First, we mentioned the use of independent assessments as a strength of the study, but it also comes with the limitation that groups of participants differ for each measurement. Such differences for which we cannot control (e.g., genetic or shared environmental differences) may also impact child mental health. To test this, we performed a supplementary mixed linear model analysis using the longitudinal data of the NTR. The results showed highly similar patterns as our main analysis and are reported in the supplementaries. Second, because of the lack of pre-covid measurements in the clinical sample, we cannot assess whether mental health problems acutely increased since the start of the pandemic as we observed in the general population, or that it took longer for the pandemic to impact the mental health of these children. Theoretically, it is also possible that mental health in the clinical sample improved at the onset of the pandemic and then returned to pre-pandemic levels, but this seems an unlikely possibility. Third, because the parent-report and self-report measures in the general population were collected in different samples, we cannot assess whether differences are due to different samples or due to different reporters, as in general correlations between raters is limited (Roy et al., 2010) and raters provide both bias and unique views (Bartels et al., 2007). Fourth, the KLIK sample may suffer from volunteer bias, the NTR sample includes relatively many participants from high SES families, and although the DREAMS sample is representative of the population in specialized child psychiatric care, low SES families are less likely to find their way into specialized care. Finally, other large-scale events such as the current war in Ukraine may have also impacted child mental health and we could not control for such effects.

## Conclusion

Internalizing child mental health problems increased during the pandemic in the general population and in children in psychiatric care. In the general population, problems were most prevalent during the first year of the pandemic, have since started to normalize, but have not yet returned to pre-pandemic levels. In the clinical population, problems increased throughout the pandemic and have not improved since. In fact, mental health problems in some domains, such as depression and sleep impairments, have worsened up until April 2022 when social restrictions were largely relaxed again. We found no evidence of sex effects and limited evidence suggesting some problems might normalize in younger children more slowly. Children from the general population seem to be more resilient to negative mental health effects of the pandemic than children in psychiatric care, in whom we see more long-term effects. We stress the importance of monitoring mental health in children throughout and beyond the pandemic to aid health care and policy.

### Key points and relevance

- Children from the general population reported increased mental and social health problems during the first year of the COVID-19 pandemic. This has since improved, but not to pre-pandemic levels.
- Children from a population in psychiatric care reported increased mental and social health problems throughout the pandemic, which have not improved up to April 2022.
- The pandemic did not impact the mental and social health of boys and girls in our samples differently.
- As there is large variability in how children reacted to the pandemic, future studies should study determinants of better and worse outcomes.
- Further changes in child mental and social health should be monitored comprehensively to aid health care and policy.

## Data Availability

All data produced in the present study are available upon reasonable request to the authors

## Acknowledgements

We thank all participating families. This research was funded by ZonMw project number 50-56300-98-973. Data collection by KLIK (measurement 1 and 2) was supported by Stichting Steun Emma Kinderziekenhuis. PROMIS reference data collection was supported by the National Health Care Institute and the Netherlands Organization for Health Research and Development. Data collection in the NTR was supported by: NWO large investment (480-15-001/674; Netherlands Twin Registry Repository: researching the interplay between genome and environment). MB is supported by a European Research Council consolidator Grant (WELL-BEING 771057 PI Bartels).

---

**Table S1.** BPM parent-report sum score estimated marginal means (EMM), standard errors, comparisons between measurement points, and % elevated scores

| Cohort |  | 0 (a)<br>pre-pandemic | 1 (b)<br>Apr/May 2020 | 2 (c)<br>Nov/Dec 2020 | 3 (d)<br>Mar/Apr 2021 | 4 (e)<br>Nov/Dec 2021 | 5 (f)<br>Mar/Apr 2022 |
| --- | --- | --- | --- | --- | --- | --- | --- |
| NTR | N | 13341 | 1332 | 221 | 347 | 426 | 458 |
|  | BPM Internalizing | 0.89 (0.01)<sup>bcdef</sup> | 1.45 (0.04)<sup>adef</sup> | 1.24 (0.11)<sup>adef</sup> | 1.77 (0.09)<sup>abc</sup> | 1.71 (0.08)<sup>abc</sup> | 1.72 (0.08)<sup>abc</sup> |
|  | BPM Externalizing | 2.05 (0.02) | 2.07 (0.06) | 2.04 (0.14) | 2.25 (0.11) | 2.15 (0.10) | 2.11 (0.10) |
|  | BPM Int % elevated | 6.9% | 16.4% | 13.5% | 19.9% | 20.2% | 19.9% |
|  | BPM Ext % elevated | 8.2% | 7.8% | 9.0% | 11.5% | 8.5% | 8.5% |
| DREAMS | N | - | 404 | 599 | 445 | 413 | 295 |
|  | BPM Internalizing | - | 5.02 (0.17)<sup>def</sup> | 5.2 (0.14)<sup>def</sup> | 5.7 (0.17)<sup>bc</sup> | 5.7 (0.17)<sup>bc</sup> | 5.69 (0.2)<sup>bc</sup> |
|  | BPM Externalizing | - | 4.5 (0.17) | 4.74 (0.15) | 4.88 (0.17) | 4.81 (0.17) | 4.83 (0.2) |
|  | BPM Int % elevated | - | 62.9% | 64.6% | 72.8% | 72.2% | 71.5% |
|  | BPM Ext % elevated | - | 38.1% | 38.9% | 39.8% | 39.7% | 40.7% |

Note. <sup>a,b,c,d,e,f</sup> represent significant differences at $p < .05$ between measurements using Least Significant Differences post-hoc tests

**Table S2.** PROMIS self-report T-score estimated marginal means (EMM), standard errors, and comparisons between measurement points

| Cohort |  | 0 (a)<br>pre-pandemic | 1 (b)<br>Apr/May 2020 | 2 (c)<br>Nov/Dec 2020 | 3 (d)<br>Mar/Apr 2021 | 4 (e)<br>Nov/Dec 2021 | 5 (f)<br>Mar/Apr 2022 |
| --- | --- | --- | --- | --- | --- | --- | --- |
| KLIK | N | 527-1082* | 471-486 | 425-440 | 407-413 | 401-414 | 514-529 |
|  | Anxiety† | 43.6 (0.3)<sup>bcdef</sup> | 50.4 (0.4)<sup>af</sup> | 50.0 (0.5)<sup>a</sup> | 50.1 (0.5)<sup>a</sup> | 50.1 (0.5)<sup>a</sup> | 49.1 (0.4)<sup>ab</sup> |
|  | Depressive symptoms† | 44.7 (0.3)<sup>bcdef</sup> | 49.3 (0.5)<sup>a</sup> | 49.0 (0.5)<sup>a</sup> | 49.9 (0.5)<sup>af</sup> | 49.5 (0.5)<sup>af</sup> | 48.2 (0.5)<sup>ade</sup> |
|  | Sleep-related impairments† | 47.0 (0.4)<sup>bcdef</sup> | 49.9 (0.5)<sup>a</sup> | 50.4 (0.5)<sup>a</sup> | 50.5 (0.5)<sup>a</sup> | 50.5 (0.5)<sup>a</sup> | 49.8 (0.4)<sup>a</sup> |
|  | Anger† | 44.1 (0.4)<sup>bcdef</sup> | 47.2 (0.5)<sup>af</sup> | 47.3 (0.5)<sup>af</sup> | 47.3 (0.5)<sup>af</sup> | 47.6 (0.5)<sup>af</sup> | 45.7 (0.4)<sup>abcde</sup> |
|  | Global health‡ | 46.6 (0.4)<sup>cd</sup> | 46.0 (0.4)<sup>d</sup> | 45.3 (0.4)<sup>a</sup> | 44.6 (0.4)<sup>abf</sup> | 45.6 (0.4) | 46.0 (0.4)<sup>d</sup> |
|  | Peer relations‡ | 47.3 (0.4)<sup>bcde</sup> | 44.5 (0.4)<sup>acdef</sup> | 46.2 (0.4)<sup>ab</sup> | 45.9 (0.4)<sup>abf</sup> | 46.2 (0.4)<sup>ab</sup> | 47.0 (0.4)<sup>bd</sup> |
| DREAMS | N | - | 241-257 | 434-456 | 236-250 | 199-216 | 169-181 |
|  | Anxiety† | - | 52.1 (0.6)<sup>cdef</sup> | 54.5 (0.5)<sup>b</sup> | 54.0 (0.6)<sup>be</sup> | 55.9 (0.6)<sup>bd</sup> | 54.3 (0.7)<sup>b</sup> |
|  | Depressive symptoms† | - | 53.1 (0.6)<sup>cdef</sup> | 55.7 (0.5)<sup>b</sup> | 56.1 (0.7)<sup>b</sup> | 55.7 (0.7)<sup>b</sup> | 56.5 (0.8)<sup>b</sup> |
|  | Sleep-related impairments† | - | 54.3 (0.6)<sup>cdef</sup> | 56.2 (0.5)<sup>b</sup> | 56.1 (0.6)<sup>b</sup> | 56.1 (0.7)<sup>b</sup> | 56.9 (0.7)<sup>b</sup> |
|  | Anger† | - | 51.0 (0.6)<sup>cdef</sup> | 53.2 (0.5)<sup>b</sup> | 53.8 (0.7)<sup>b</sup> | 53.5 (0.7)<sup>b</sup> | 53.5 (0.8)<sup>b</sup> |
|  | Global health‡ | - | 42.2 (0.5)<sup>cdef</sup> | 40.7 (0.4)<sup>bd</sup> | 38.9 (0.5)<sup>bc</sup> | 40.0 (0.6)<sup>b</sup> | 39.5 (0.6)<sup>b</sup> |
|  | Peer relations‡ | - | 43.7 (0.5) | 44.4 (0.4)<sup>de</sup> | 42.3 (0.6)<sup>c</sup> | 43.5 (0.6) | 42.6 (0.7)<sup>c</sup> |

Note. <sup>a,b,c,d,e,f</sup> represent significant differences at $p < .05$ between measurements using Least Significant Differences post-hoc tests.
\* Sample sizes vary because data from different domains comes from different norm studies.
† Higher scores indicate more symptoms
‡ Higher scores indicate better functioning

**Table S3.** PROMIS % normal, moderately elevated, and severely elevated scores

|  |  | KLIK |  |  |  |  |  | DREAMS |  |  |  |  |  |
| --- | --- | --- | --- | --- | --- | --- | --- | --- | --- | --- | --- | --- | --- |
|  |  | 0 (a)<br>pre-pandemic | 1 (b)<br>Apr/May<br>2020 | 2 (c)<br>Nov/Dec<br>2020 | 3 (d)<br>Mar/Apr<br>2021 | 4 (e)<br>Nov/Dec<br>2021 | 5 (f)<br>Mar/Apr<br>2022 | 0 (a)<br>pre-<br>pandemi<br>c | 1 (b)<br>Apr/May<br>2020 | 2 (c)<br>Nov/Dec<br>2020 | 3 (d)<br>Mar/Apr<br>2021 | 4 (e)<br>Nov/Dec<br>2021 | 5 (f)<br>Mar/Apr<br>2022 |
| N |  | 527-1082* | 471-486 | 425-440 | 407-413 | 401-414 | 514-529 | - | 241-257 | 434-456 | 236-250 | 199-216 | 169-181 |
| Anxiety | normal | 74.8% | 46.6% | 50.9% | 51.0% | 52.9% | 56.4% | - | 45.2% | 32.7% | 35.7% | 29.7% | 36.9% |
|  | moderate | 20.0% | 49.3% | 43.1% | 39.0% | 37.3% | 35.4% | - | 40.8% | 43.3% | 49.2% | 45.8% | 42.6% |
|  | severe | 5.2% | 4.2% | 6.0% | 10.0% | 9.8% | 8.2% | - | 14.0% | 24.0% | 15.2% | 24.5% | 20.5% |
| Depressive | normal | 74.8% | 60.0% | 61.2% | 55.5% | 60.6% | 63.8% | - | 52.2% | 38.6% | 38.8% | 39.5% | 34.1% |
| Symptoms | moderate | 20.1% | 37.2% | 33.2% | 37.9% | 32.2% | 31.0% | - | 31.0% | 32.9% | 37.6% | 41.0% | 38.2% |
|  | severe | 5.1% | 2.7% | 5.6% | 6.6% | 7.2% | 5.2% | - | 16.7% | 28.5% | 23.6% | 19.5% | 27.7% |
| Sleep-related | normal | 71.5% | 64.5% | 58.2% | 55.4% | 55.9% | 61.8% | - | 50.2% | 39.2% | 39.8% | 39.8% | 36.0% |
| impairments | moderate | 23.3% | 33.4% | 37.3% | 40.0% | 39.4% | 34.9% | - | 38.7% | 44.7% | 47.0% | 46.3% | 43.6% |
|  | severe | 5.1% | 2.1% | 4.5% | 4.7% | 4.7% | 3.3% | - | 11.1% | 16.1% | 13.1% | 13.9% | 20.3% |
| Anger | normal | 75.0% | 73.5% | 69.6% | 68.5% | 68.0% | 75.9% | - | 54.9% | 46.7% | 41.5% | 45.9% | 45.1% |
|  | moderate | 19.9% | 23.6% | 25.3% | 24.6% | 24.9% | 20.7% | - | 28.9% | 35.1% | 42.3% | 37.3% | 39.4% |
|  | severe | 5.1% | 2.9% | 5.1% | 6.8% | 7.1% | 3.4% | - | 16.3% | 18.1% | 16.2% | 16.7% | 15.4% |
| Global | normal | 75.0% | 72.0% | 66.8% | 66.3% | 68.8% | 71.3% | - | 59.9% | 44.3% | 41.2% | 46.8% | 39.2% |
| Health | moderate | 20.1% | 26.3% | 27.7% | 27.8% | 25.4% | 23.4% | - | 27.2% | 29.6% | 34.0% | 31.0% | 31.5% |
|  | severe | 5.0% | 1.6% | 5.5% | 5.8% | 5.8% | 5.3% | - | 12.8% | 26.1% | 24.8% | 22.2% | 29.3% |
| Peer | normal | 74.2% | 71.3% | 77.2% | 75.4% | 76.8% | 79.6% | - | 66.0% | 68.7% | 55.5% | 63.3% | 60.9% |
| relations | moderate | 19.9% | 22.9% | 17.4% | 19.7% | 17.7% | 14.8% | - | 22.4% | 23.0% | 34.3% | 28.6% | 29.0% |
|  | severe | 5.9% | 5.7% | 5.4% | 4.9% | 5.5% | 5.6% | - | 11.6% | 8.3% | 10.2% | 8.0% | 10.1% |

\* Sample sizes vary because data from different domains comes from different norm studies.

## Supplementary analyses

For the NTR general population, within-person data exists for all measurement moments, including the pre-covid measurements. Therefore, we modeled a pre-pandemic versus during covid variable, a variable representing all COVID measurements, and a variable representing the quadratic effect of all COVID measurements. In addition, we included age, sex, and interactions between time-variables and age and sex.

We carried out the linear mixed model analysis using nlme package (version 3.1-159) in R (version 4.1.0). We estimated the model using a maximum likelihood estimator. The following equation describes the model: Y_ij_ = (β_1_ + b_1i_) + (β_2_ + b_2i_) * Cov + (β_3_ + b_3i_) * time + (β_4_ +b_4i_) * time^2^ + β_5_ * sex + β_6_ * age + β_7_ * Cov * sex + β_8_ * Cov * age + β_9_ * time * sex + β_10_ * time * age + e_ij_, where Y_ij_ is the internalizing/externalizing score for individual i at measurement j, Cov is the variable specifying a measurement during the pandemic, e_ij_ is the error term, β1, …, β10 are the fixed effects (population averages) and b_1i_, …, b_4i_ are the individual specific random intercept and slopes.

We observed a significant increase in internalizing scores from pre-pandemic to pandemic measurements (t = 3.46, p < .001), with no significant changes in subsequent measurements. There was a significant interaction between pre-pandemic versus pandemic and age (t = −2.09, p < .05) with younger children showing larger increases in internalizing problems, but no significant interaction with sex. There was a significant interaction between time during COVID and sex (t = 1.98, p < .05), with girls having higher internalizing problems over time, but no significant interaction with age.

We observed a significant increase in externalizing scores from pre-pandemic to pandemic measurements (t = −2.86, p < .01), with no significant changes in subsequent measurements. There were no significant interactions between start of covid lockdown measures and age or sex, nor between time during covid and age, but there was a significant interaction between time during covid and sex (t = 2.58, p < 05).

